# Ferritin across long-term conditions in England: cross-sectional primary care study

**DOI:** 10.64898/2026.06.06.26355042

**Authors:** A Katumba, C Wright Drakesmith, S Haynes, S Maynard, V Maharajan, I Erone, M Smith, A Shah, N Roy, C Bankhead, SJ Stanworth

**Affiliations:** Nuffield Department of Primary Care Health Sciences, University of Oxford, UK; NIHR Blood and Transplant Research Unit in Data Driven Transfusion Practice, Radcliffe Department of Medicine, University of Oxford, UK; Department of Haematology, University Hospitals NHS Foundation Trust, Oxford, UK; Department of Haematology/Transfusion Medicine, NHS Blood and Transplant, John Radcliffe Hospital, Oxford University Hospitals NHS Trust, UK; Nuffield Department of Women’s and Reproductive Health, University of Oxford, UK; Radcliffe Department of Medicine, John Radcliffe Hospital, University of Oxford, UK; Department of Anaesthesia, Hammersmith Hospital, Imperial College Healthcare NHS Trust, London, UK

**Keywords:** Ferritin, Iron Deficiency, Primary Health Care, Chronic Disease, Epidemiology

## Abstract

**Background:** Iron deficiency (ID) is a readily treatable condition once identified. Ferritin is the primary diagnostic marker, but cut-offs vary and inflammation complicates interpretation in patients with long-term conditions (LTCs).

**Aim:** To describe ferritin distribution and the prevalence of threshold-defined low ferritin in adults with and without LTCs in primary care.

**Design and setting:** Cross-sectional observational study using routinely collected electronic health records from a national primary care database in England (1^st^ January 2015 to 31^st^ December 2021).

**Method:** Adults with ≥1 ferritin test in Clinical Practice Research Datalink (CPRD) Aurum were included. LTCs were identified using validated primary-care code lists. Outcomes included ferritin distribution and threshold-defined ID prevalence using World Health Organization (WHO) (<15 µg/L; <70 µg/L if inflammation) and National Institute for Health and Care Excellence (NICE) (<30 µg/L) cut-offs, stratified by sex and, in women, by age <50 versus ≥50 as a proxy for menopausal status.

**Results:** 4,489,594 individuals were included; 55% (n=2,469,882) had ≥1 LTC. Ferritin was lowest in women <50 and in LTCs characterised by impaired absorption or blood loss (coeliac disease, inflammatory bowel disease). Among women <50 with an LTC, 80% had ferritin <70 µg/L versus 47% <30 µg/L, leaving 33% in the 30–70 µg/L range potentially missed by standard cut-offs; equivalent figures were 28% in women ≥50 and 17% in men.

**Conclusion:** Threshold-defined low ferritin is very common across LTCs and disproportionately affects women, particularly those under 50. Condition-specific, inflammation-adjusted ferritin thresholds may improve detection, management, and equity in primary care.

## Introduction

Iron deficiency (ID) is a major cause of morbidity, contributing to reduced physical performance, reduced work productivity, increased hospitalisation rates and higher mortality (1–8). ID, whether associated with anaemia or not, is commonly tested for in patients with inflammatory co-morbidities, or long-term conditions (LTCs), given that ID is common in LTCs and associated with poorer clinical outcomes (9–13).

Ferritin is the most widely used biomarker of iron status, however the threshold for defining possible ID remains contested. The World Health Organization (WHO) recommends a cut-off of <15 μg/L, while the UK National Institute for Health and Care Excellence (NICE) suggest a threshold of <30 μg/L (14). Interpreting ferritin is further complicated by its role as an acute-phase reactant. In individuals with LTCs, inflammation can elevate ferritin independent of iron stores, masking underlying ID, therefore delaying recognition and treatment(15). To address this, higher ferritin thresholds have been proposed in selected inflammatory states. A threshold of 100μg/L is recommended for patients with chronic kidney disease (CKD) and heart failure by NICE and the European Society of Cardiology respectively (16,17). The WHO suggests a threshold of <70 μg/L in the presence of inflammation (14), however does not specify which conditions warrant this adjustment, leading to uncertainty and inconsistent application in clinical practice. Applying <70 μg/L uniformly across LTCs is therefore a pragmatic descriptive approach and does not imply equivalent inflammatory burden across conditions.

This study describes the distribution of ferritin and the prevalence of threshold-defined low ferritin among patients with and without LTCs using large-scale UK primary care data from Clinical Practice Research Datalink (CPRD) Aurum. This study aims to inform future evaluation of whether stratified thresholds might improve identification and management of possible ID in patients with LTCs.

## Methods

We conducted a retrospective cross-sectional observational study using anonymised patient data from the CPRD Aurum database, a UK primary care database covering 41 million patients across 1,489 practices. We included 4,489,594 individuals with at least one ferritin test.

### Study Population

Adults aged ≥18 years with at least one ferritin test between January 1, 2015, and December 31, 2021, were included. The study population included patients with and without LTCs prior to study entry. LTCs are defined as conditions of long duration requiring ongoing clinical management. Included LTCs were selected by the research team based on prevalence within the UK healthcare system, potential association with inflammation, and alignment with core conditions identified through expert Delphi consensus for inclusion in multimorbidity research (18).

LTCs included bronchiectasis, cirrhosis, chronic kidney disease (CKD) stages 3–5, chronic obstructive pulmonary disease (COPD), dementia, heart failure, multiple sclerosis, obesity, osteoarthritis, Parkinson’s disease, rheumatoid arthritis, stroke and type 1 and type 2 diabetes mellitus. We also included conditions more typically associated with local inflammation, impaired absorption, or increased iron loss; such as coeliac disease, Crohn’s disease, ulcerative colitis, gastroesophageal reflux disease (GORD), polycystic ovary syndrome (PCOS) and endometriosis.

### Data Collection

Demographic data and clinical parameters were extracted. Threshold-defined low ferritin was defined using WHO suggested thresholds (ferritin <15 µg/L for apparently healthy individuals and <70 µg/L for patients with inflammation) and NICE suggested thresholds (ferritin <30 µg/L). Clinician validated SNOMED-CT code lists were used to identify LTCs. Codelists used for data extraction are available at (22-001873-Anaemia/Katumba at main · ndpchs-cprd/22-001873-Anaemia · GitHub).

### Outcome Measures

Primary outcomes were:

1. Ferritin level distribution
2. Prevalence of threshold-defined low ferritin, defined as at least one ferritin result meeting the WHO or NICE threshold

### Statistical Analysis

Descriptive statistics summarised results. Age was categorised into three groups (men; women <50 years; women ≥50 years) to reflect physiologically meaningful differences in iron metabolism, particularly the shift in menstrual blood loss and iron requirements around the menopause. Where patients had multiple ferritin tests, the lowest ferritin result per person was used for both ferritin distribution and threshold-based analyses.

Ferritin concentrations were log_2_-transformed to reduce skewness, stabilise variance and allow fold-change interpretation; percentiles were back-transformed to µg/L for clinical interpretability. Ferritin distributions were summarised using the median and interquartile range (25th–75th percentiles), with whiskers representing the 2.5th–97.5th percentiles. Threshold-based proportions are reported with binomial (Wilson) 95% confidence intervals.

Given the large sample size (n≈4.5 million), formal hypothesis tests would detect statistically significant differences for nearly all comparisons regardless of clinical magnitude. We therefore adopted an estimation approach throughout, interpreting differences on the basis of effect size, confidence interval width, and clinical relevance rather than p-values, consistent with recognised limitations of null hypothesis significance testing in large samples (19,20). For multimorbidity comparisons (Supplementary Tables S6a–b), ferritin is additionally summarised as geometric means with 95% CIs to facilitate comparison between single-LTC and multiple-LTC groups. All analyses were conducted in R (RStudio).

## Results

### Cohort Characteristics

The study cohort included 4,489,594 individuals with at least one ferritin test (Table 1). Of these, 65% were female (n=2,924,273), 55% had at least one LTC (n=2,469,882) and 57% had only one ferritin test (n=2,543,585). The percentage of individuals with at least one LTC increased with age, reaching 85% in the 80+ age group. In total, 8,774,103 ferritin tests were performed with 68% (n=5,958,941) performed in women (Supplementary Table S1-2).

**Table 1.** Characteristics of the study population by LTC status. Counts are presented as n (), stratified by LTC status.

| Characteristic | Total, n(%) | LTC Present, n(%) | No LTC, (%) |
| --- | --- | --- | --- |
| **Total individuals N** | 4489594 (100) | 2469882 (55) | 2019712 (45) |
| **Gender** |  |  |  |
| F | 2924273 (65.1) | 1577748 (54) | 1346525 (46) |
| M | 1565321 (34.9) | 892134 (57) | 673187 (43) |
| **Age category** |  |  |  |
| 18-39 | 1456134 (32.4) | 446890 (30.7) | 1009244 (69.3) |
| 40-49 | 781255 (17.4) | 381627 (48.8) | 399628 (51.2) |
| 50-59 | 786394 (17.5) | 471093 (59.9) | 315301 (40.1) |
| 60-69 | 659358 (14.7) | 466055 (70.7) | 193303 (29.3) |
| 70-79 | 630603 (14) | 498185 (79) | 132418 (21) |
| 80+ | 553774 (12.3) | 470024 (84.9) | 83750 (15.1) |
| **Ethnicity category** |  |  |  |
| Asian, Asian British or Asian Welsh | 454564 (10) | 218255 (48) | 236309 (52) |
| Black, Black British, Black Welsh, Caribbean or African | 206076 (5) | 113991 (55) | 92085 (45) |
| Mixed or Multiple ethnic groups | 68913 (2) | 29960 (43) | 38953 (57) |
| Other ethnic group | 67058 (1) | 26948 (40) | 40110 (60) |
| Unknown | 531032 (12) | 252472 (48) | 278560 (52) |
| White | 3161951 (70) | 1828256 (58) | 1333695 (42) |
| **Number of ferritin tests** |  |  |  |
| 1 | 2543585 (56.7) | 1234439 (48.5) | 1309146 (51.5) |
| 2 | 963621 (21.5) | 562121 (58.3) | 401500 (41.7) |
| 3 | 445314 (9.9) | 286885 (64.4) | 158429 (35.6) |
| 4 | 229550 (5.1) | 157618 (68.7) | 71932 (31.3) |
| 5 | 125193 (2.8) | 89787 (71.7) | 35406 (28.3) |
| 6-10 | 163371 (3.6) | 123680 (75.7) | 39691 (24.3) |
| 11-20 | 17577 (0.4) | 14187 (80.7) | 3390 (19.3) |
| 21+ | 1383 (0) | 1165 (84.2) | 218 (15.8) |

### Ferritin Levels

Conditions associated with GI inflammation, malabsorption or blood loss (coeliac disease, Crohn’s disease, ulcerative colitis) consistently showed the lowest median ferritin across all groups, while no single condition category consistently showed the highest values. Median ferritin varied more across conditions in men (70–122 µg/L) than in women under 50 (27–45 µg/L). T2DM had the lowest median ferritin across every stratum.

### Low ferritin using different guideline thresholds

Figure 2 highlights the prevalence of threshold-defined low ferritin stratified by LTC status and sex using three ferritin thresholds: <15 µg/L (WHO), <30 µg/L (NICE), and <70 µg/L (WHO, inflammation-adjusted). Women consistently had a higher prevalence of threshold-defined low ferritin than men at all thresholds. Among both men and women, the overall prevalence at <15 µg/L and <30 µg/L was broadly similar between those without and those with LTCs.

**Figure 1.**
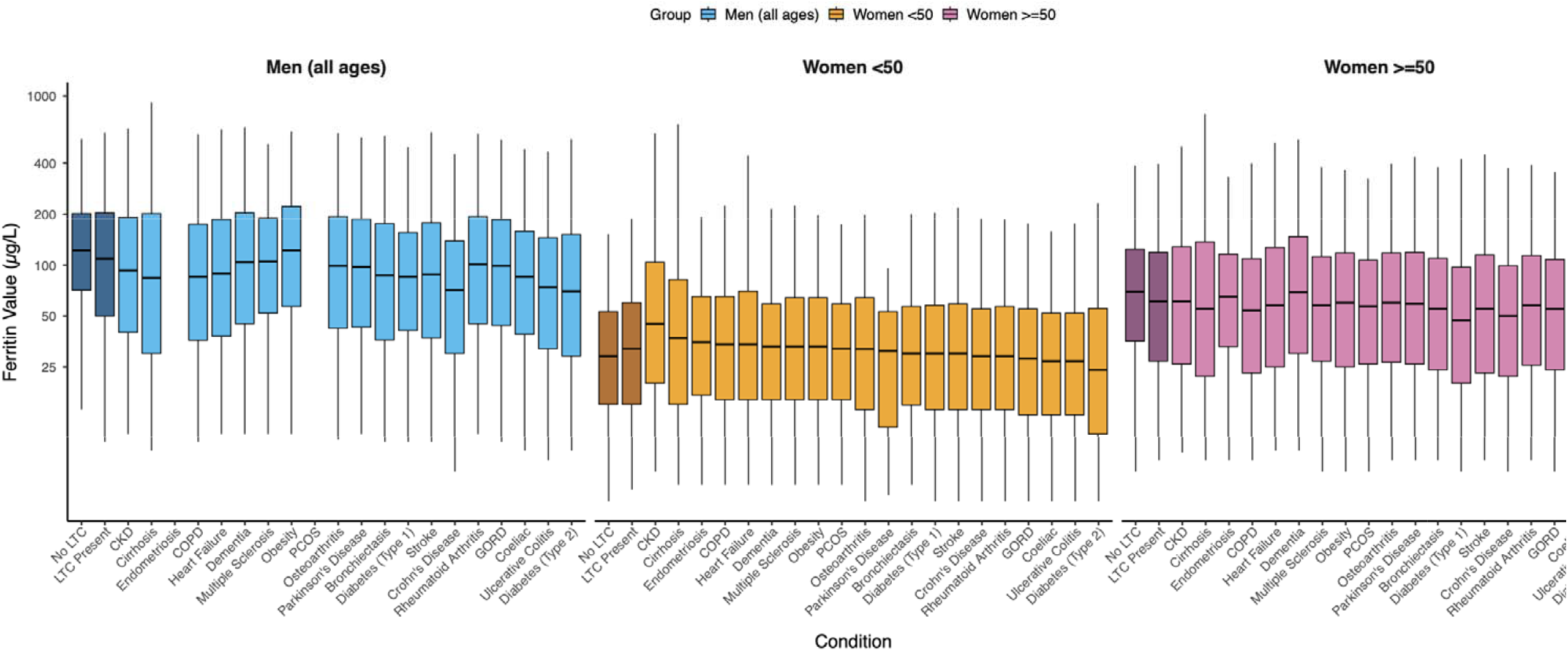
Ferritin levels by long-term condition (LTC) status and sex. Distribution of the lowest recorded serum ferritin concentration (µg/L) by LTC status and sex (men; women <50 years; women ≥50 years). Values were analysed on the log_2_ scale and are presented on the original scale for interpretability. The central line represents the median; the box the interquartile range (25th–75th percentiles); whiskers extend to the 2.5th and 97.5th percentiles.

**Figure 2.**
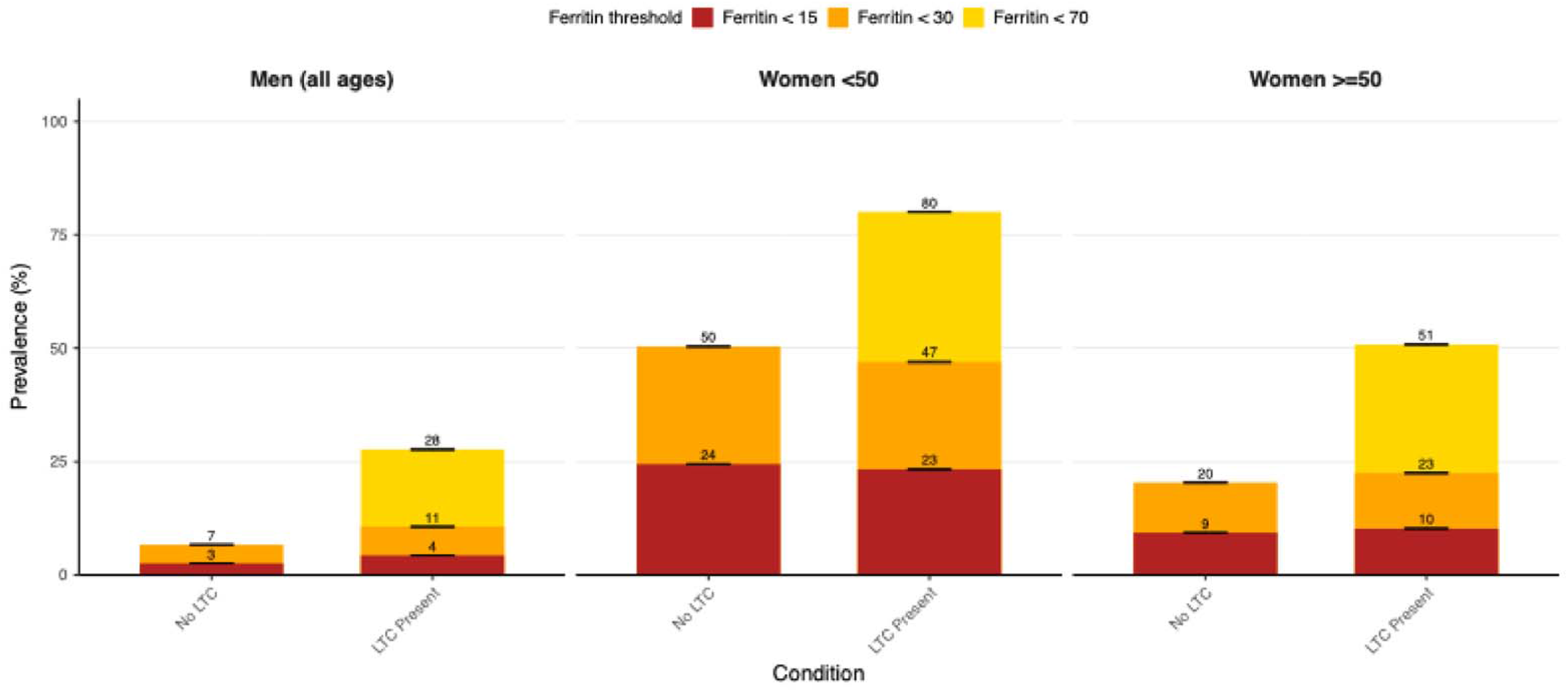
Prevalence of threshold-defined low ferritin by LTC status and sex (<15, <30, and <70 µg/L). Bars show the proportion (%) of individuals whose lowest recorded ferritin fell below each threshold, stratified by LTC status and sex; whiskers indicate binomial 95% confidence intervals. The shaded band between <30 and <70 µg/L highlights individuals who meet the WHO inflammation-adjusted threshold but would not be identified using the standard NICE cut-off.

Among women under 50 with at least one LTC, 80% had ferritin <70 µg/L compared with 47% below 30 µg/L, meaning 33% fell in the 30–70 µg/L range potentially overlooked if only standard NICE thresholds are applied. In women aged 50 and over with LTCs, 51% had ferritin <70 µg/L versus 23% below 30 µg/L, giving a similar 28% in this intermediate range. A less pronounced but consistent pattern was seen in men, where 28% of those with LTCs had ferritin <70 µg/L compared with 11% below 30 µg/L, leaving 17% in the 30–70 µg/L range.

### Low ferritin by LTC

Figure 3 illustrates the prevalence of threshold-defined low ferritin across LTC cohorts using the WHO ferritin thresholds to allow descriptive inflammation-adjusted comparisons across conditions. Inflammatory bowel diseases (IBDs; including Crohn’s disease and ulcerative colitis) and coeliac disease were consistently associated with higher rates of threshold-defined low ferritin. Both T1DM and T2DM also showed a strong association with threshold-defined low ferritin, particularly in women.

**Figure 3.**
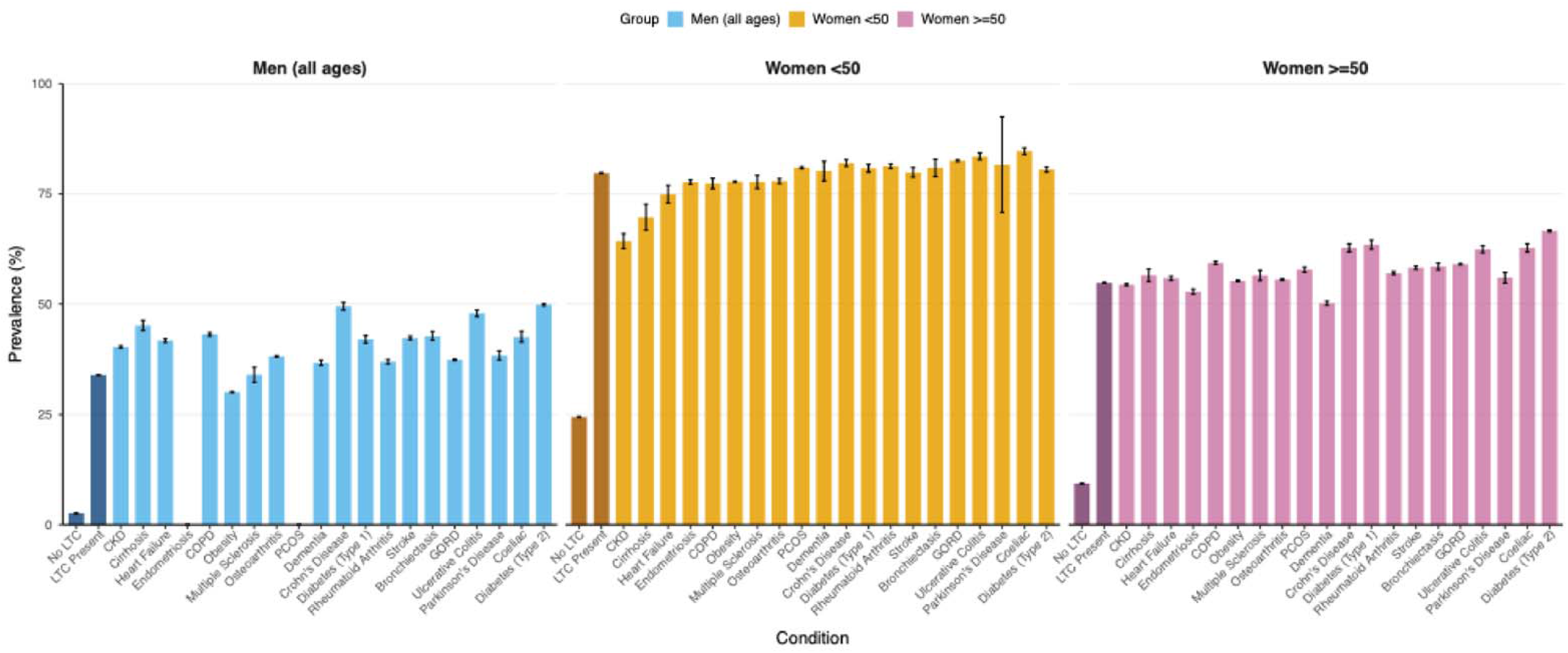
Prevalence of threshold-defined low ferritin by LTC status and sex, applying WHO thresholds (<15 µg/L in those without LTCs; <70 µg/L in those with LTCs). Bars show the proportion (%) of individuals whose lowest recorded ferritin fell below the applicable threshold, stratified by sex (men; women <50 years; women ≥50 years); whiskers indicate binomial 95% confidence intervals.

### Multiple Long-Term Conditions (MLTCs)

Men with MLTCs had a higher prevalence of threshold-defined low ferritin than those with a single LTC across virtually all conditions, most markedly in obesity (19.5% rising to 38.3%; Supplementary Table S6b). IBD was an exception, with little additional burden above the already high single-LTC prevalence. In women, the effect of multimorbidity differed by age. Women under 50 with MLTCs had marginally lower prevalence of threshold-defined low ferritin compared to those with single LTCs, while women aged 50 and over with MLTCs had consistently higher prevalence of threshold-defined low ferritin across nearly all conditions (Supplementary Tables S6a–b).

## Discussion

Ferritin levels and threshold-defined low ferritin prevalence were found to vary by sex, age, and multimorbidity. Conditions associated with GI inflammation, malabsorption or blood loss (coeliac disease and IBD) consistently showed the lowest ferritin across all strata. CKD showed notably elevated values particularly in women, consistent with inflammation-driven ferritin elevation rather than true iron repletion. The lower ferritin in women under 50 compared with older women and men is consistent with menstrual iron losses ceasing around the menopause. In men, ferritin varied widely across conditions (70–122 µg/L), whereas in women under 50 most conditions fell within a narrow range (27–45 µg/L). This narrower range suggests that menstrual iron loss is the dominant driver of ferritin in younger women, overriding condition-specific effects that are more apparent in men. These patterns support evaluation of whether condition-stratified thresholds could improve identification of possible ID.

High rates of threshold-defined low ferritin were observed across several conditions not routinely screened in clinical practice, including PCOS, GORD, T1DM and T2DM (21–24). T2DM had the highest prevalence of threshold-defined low ferritin of all conditions in men and in women aged 50 and over, suggesting drivers of possible ID in diabetes that merit further investigation. Together, these findings highlight a broader gap between epidemiological evidence and clinical practice across several LTC groups that warrants further attention during future guideline development.

Most importantly, we observed a diagnostic gap between general and inflammation-aware thresholds. Among women under 50 with at least one LTC, 80% had ferritin <70 µg/L compared with 47% below 30 µg/L, leaving approximately one-third (33%) in the 30–70 µg/L range that would be missed if only standard NICE thresholds are applied. In women aged 50 and over with LTCs, 51% had ferritin <70 µg/L versus 23% below 30 µg/L, leaving a similar 28% in this intermediate range. In men, 17% fell in the 30–70 µg/L range. This represents a meaningful population at risk of undetected iron depletion, warranting clinical review in the context of each individual’s symptoms and additional iron markers. The proportional impact is greatest in younger women. Together, these findings suggest that a single universal threshold risks misclassification in LTC settings, particularly in women, and support further evaluation of condition-stratified ferritin thresholds.

Multimorbidity amplified threshold-defined low ferritin prevalence in men across nearly all conditions, most strikingly in obesity where prevalence nearly doubled (19.5% to 38.3%). IBD was an exception, with little additional burden, consistent with prevalence already being near ceiling in single-LTC IBD. In women, the effect of multimorbidity differed by age. Among women under 50, threshold-defined low ferritin prevalence was marginally lower with MLTCs, possibly reflecting compositional shifts in menstrual status; in women aged 50 and over, multimorbidity consistently increased threshold-defined low ferritin prevalence, converging with the male pattern and suggesting that comorbidity burden becomes the dominant driver of low ferritin risk once menstruation ceases.

### Strengths and Limitations

The study’s strength lies in its large, real-world dataset, enabling detailed description of ferritin distributions and threshold-defined low ferritin. CPRD is broadly representative of the UK population by age and sex, and its granular stratification by age, sex, and comorbidities allows a nuanced assessment of ferritin variability across cohorts (25).

Despite these strengths, the study has several limitations. First, the analysis lacked hospital laboratory data, limiting generalisability beyond primary care and potentially under-recording ferritin tests undertaken in secondary care (for example specialist clinics). Reliance on primary care diagnostic coding may also have misclassified some individuals as having “no LTC” if diagnoses were recorded solely in secondary care.

Second, selection bias is a major limitation because inclusion required at least one recorded ferritin test and individuals tested in routine practice are unlikely to represent the underlying population. Ferritin is typically requested for suspected iron deficiency anaemia, monitoring iron therapy, investigation of suspected iron overload, or at patient request. Therefore, the observed distributions and proportions with low ferritin likely overestimate, rather than underestimate, the prevalence in the untested primary care population and should not be treated as general population estimates.

Residual confounding is also possible. We did not measure antiplatelet or anticoagulant prescribing, which may increase bleeding risk and affect both ferritin levels and the likelihood of testing. In women, menopausal status is not directly recorded in CPRD. Although age-stratified analyses were performed using prespecified life-stage bands, these are only an imperfect proxy for menopausal status and cannot capture individual variation in menstrual status, hysterectomy, hormone therapy, or heavy menstrual bleeding. Female subgroup comparisons should therefore be interpreted cautiously.

Finally, although ferritin is the most commonly used test to indicate iron deficiency, it is imprecise and not the gold standard (26). Sparse measurement and non-contemporaneous availability of transferrin saturation, total iron binding capacity and C-reactive protein (CRP) in primary care limits the ability to distinguish true iron deficiency from inflammation-driven changes. These limitations reflect the nature of real-world clinical data not collected for research purposes. Future work using cohorts with systematic biomarker measurement independent of clinical indication (for example UK Biobank) could better support population-level prevalence estimates, while recognising that such cohorts also have their own selection characteristics.

### Comparison with Existing Literature

Our findings align with reports of higher ferritin in heart failure and CKD, and lower ferritin with higher prevalence of threshold-defined low ferritin in coeliac disease and IBDs(16,27,28). Sex differences also reflect existing evidence of greater ID risk in women (22). Our study adds novel insights by demonstrating that the gap between standard and inflammation-adjusted thresholds is substantially larger in women, particularly those under 50, and that several conditions not currently prioritised for ID screening, including PCOS, GORD, and diabetes, show high rates of threshold-defined low ferritin (16).

### Implications for Future Research

Future research should explore and define condition-specific ferritin reference ranges for patients with LTCs, as current generalised thresholds risk underdiagnosis due to inflammation-related ferritin elevation. Broader biomarker panels (e.g. CRP, ESR, MCV) and concurrent testing could help distinguish true ID from inflammation, particularly in CKD, heart failure, and cirrhosis. Replication of these findings in international cohorts is needed to assess generalisability and explore the influence of healthcare practice and policy.

### Implications for Clinical Practice

Several conditions, including IBD, coeliac disease, diabetes, PCOS, and GORD, show substantial proportions of patients with ferritin in the 30-70 µg/L range, potentially missed by standard NICE thresholds. These groups may warrant consideration for expanded ferritin screening, though ferritin alone cannot confirm iron deficiency and clinical assessment remains necessary before any treatment decisions. Updating guidelines to consider ferritin testing in these under-recognised groups may support early detection and reduce diagnostic delays.

## Conclusion

This study highlights marked variation in ferritin distributions across LTC and demographic groups, underscoring the possible need for tailored thresholds. High rates of threshold-defined low ferritin were observed across a range of conditions, including IBD, coeliac disease, T1DM, T2DM, PCOS, and GORD, several of which are not currently prioritised for routine ID screening. Existing guidelines may not fully account for the diagnostic uncertainty created by inflammation in these populations, potentially contributing to missed cases in the 30– 70 µg/L range. Establishing condition-specific ferritin reference ranges for identifying possible ID in the context of inflammation could improve clinical interpretation and reduce reliance on a single universal threshold. Electronic health records could flag ferritin in the 30–70 µg/L range in relevant LTCs, prompting clinical review. Furthermore, integrating updated evidence-based cut-offs into national guidance, such as the Quality and Outcomes Framework, could facilitate earlier identification and more consistent management of possible ID in patients with LTCs.

## Data Availability

Data may be obtained from a third party and are not publicly available. The data used in this study were obtained from the Clinical Practice Research Datalink under licence from the Medicines and Healthcare products Regulatory Agency. The individual-level data cannot be shared by the authors because access is subject to Clinical Practice Research Datalink Research Data Governance approval, data-sharing agreements and licensing conditions. Researchers may apply for access to Clinical Practice Research Datalink data through the Clinical Practice Research Datalink Research Data Governance process. Summary data generated by this study are provided in the manuscript and supplementary materials.

## Acknowledgements

This study is based in part on data from the Clinical Practice Research Datalink obtained under licence from the UK Medicines and Healthcare products Regulatory Agency. The data is provided by patients and collected by the NHS as part of their care and support. The interpretation and conclusions contained in this study are those of the author/s alone.

We thank the participating general practices contributing to CPRD Aurum and colleagues (including Professor Hal Drakesmith) who supported analysis and interpretation.

## Notes

**<u>How this fits in</u>** Iron deficiency (ID) in patients with long-term conditions can be missed when general-population ferritin thresholds are applied in inflammatory states. Using primary care data, we show sex- and condition-specific differences in ferritin distribution and threshold-defined ID. Our findings support evaluation of condition-specific, inflammation-adjusted ferritin thresholds to improve detection of possible ID in primary care.

### Competing Interest Statement

The authors have declared no competing interest.

### Author Declarations

The Clinical Practice Research Datalink Research Data Governance process of the Medicines and Healthcare products Regulatory Agency gave ethical approval for this work (protocol number 22_001873).

## References

1. Camaschella C. Iron deficiency. Blood. 2019 Jan 3;133(1):30–9. doi:10.1182/blood-2018-05-815944

2. Stahl-Gugger A, de Godoi Rezende Costa Molino C, Wieczorek M, Chocano-Bedoya PO, Abderhalden LA, Schaer DJ, et al. Prevalence and incidence of iron deficiency in European community-dwelling older adults: an observational analysis of the DO-HEALTH trial. Aging Clin Exp Res. 2022 Sep;34(9):2205–15. doi:10.1007/s40520-022-02093-0 PubMed PMID: 35304704; PubMed Central PMCID: PMC9464157.

3. Levi M, Rosselli M, Simonetti M, Brignoli O, Cancian M, Masotti A, et al. Epidemiology of iron deficiency anaemia in four European countries: a population-based study in primary care. Eur J Haematol. 2016;97(6):583–93. doi:10.1111/ejh.12776

4. Pasricha SR, Tye-Din J, Muckenthaler MU, Swinkels DW. Iron deficiency. The Lancet. 2021 Jan 16;397(10270):233–48. doi:10.1016/S0140-6736(20)32594-0

5. Smith M, Drakesmith CW, Haynes S, Maynard S, Shah A, Roy NB, et al. Prevalence and patterns of testing for anaemia in primary care in England: a cohort study using an electronic health records database. Br J Gen Pract J R Coll Gen Pract. 2025 Apr;75(753):e232–40. doi:10.3399/BJGP.2024.0336 PubMed PMID: 39658076; PubMed Central PMCID: PMC11881008.

6. Rohr M, Brandenburg V, Brunner-La Rocca HP. How to diagnose iron deficiency in chronic disease: A review of current methods and potential marker for the outcome. Eur J Med Res. 2023 Jan 9;28(1):15. doi:10.1186/s40001-022-00922-6

7. Georgieff MK. Iron deficiency in pregnancy. Am J Obstet Gynecol. 2020 Oct;223(4):516–24. doi:10.1016/j.ajog.2020.03.006 PubMed PMID: 32184147; PubMed Central PMCID: PMC7492370.

8. Lee HS, Chao HH, Huang WT, Chen SCC, Yang HY. Psychiatric disorders risk in patients with iron deficiency anemia and association with iron supplementation medications: a nationwide database analysis. BMC Psychiatry. 2020 May 11;20(1):216. doi:10.1186/s12888-020-02621-0

9. Cacoub P, Choukroun G, Cohen-Solal A, Luporsi E, Peyrin-Biroulet L, Peoc’h K, et al. Towards a Common Definition for the Diagnosis of Iron Deficiency in Chronic Inflammatory Diseases. Nutrients. 2022 Feb 28;14(5). doi:10.3390/nu14051039 PubMed PMID: 35268014; PubMed Central PMCID: PMC8912638.

10. Cappellini MD, Comin-Colet J, de Francisco A, Dignass A, Doehner W, Lam CS, et al. Iron deficiency across chronic inflammatory conditions: International expert opinion on definition, diagnosis, and management. Am J Hematol. 2017;92(10):1068–78. doi:10.1002/ajh.24820

11. Dignass A, Farrag K, Stein J. Limitations of Serum Ferritin in Diagnosing Iron Deficiency in Inflammatory Conditions. Int J Chronic Dis. 2018 Jan 1;2018(1):9394060. doi:10.1155/2018/9394060

12. Martens P, Nijst P, Verbrugge FH, Smeets K, Dupont M, Mullens W. Impact of iron deficiency on exercise capacity and outcome in heart failure with reduced, mid-range and preserved ejection fraction. Acta Cardiol. 2018 Mar 4;73(2):115–23. doi:10.1080/00015385.2017.1351239

13. Thum T, Anker SD. Nutritional iron deficiency in patients with chronic illnesses. The Lancet. 2007 Dec 8;370(9603):1906. doi:10.1016/S0140-6736(07)61810-8

14. World Health Organization. WHO guideline on use of ferritin concentrations to assess iron status in individuals and populations [Internet]. 2020. Available from: https://www.ncbi.nlm.nih.gov/books/NBK569880/

15. Fertrin KY. Diagnosis and management of iron deficiency in chronic inflammatory conditions (CIC): is too little iron making your patient sick? Hematology. 2020 Dec 4;2020(1):478–86. doi:10.1182/hematology.2020000132

16. McDonagh TA, Metra M, Adamo M, Gardner RS, Baumbach A, Böhm M, et al. 2021 ESC Guidelines for the diagnosis and treatment of acute and chronic heart failure: Developed by the Task Force for the diagnosis and treatment of acute and chronic heart failure of the European Society of Cardiology (ESC) With the special contribution of the Heart Failure Association (HFA) of the ESC. Eur Heart J. 2021 Sep 21;42(36):3599–726. doi:10.1093/eurheartj/ehab368

17. National Institute for Health and Care Excellence. Anaemia iron deficiency: What investigations should I arrange to confirm iron deficiency anaemia?

18. Iris S S Ho, Amaya Azcoaga-Lorenzo, Ashley Akbari, Jim Davies, Kamlesh Khunti, Umesh T Kadam, et al. Measuring multimorbidity in research: Delphi consensus study. BMJ Med. 2022 Jul 27;1(1):e000247. doi:10.1136/bmjmed-2022-000247

19. Wasserstein RL, Lazar NA. The ASA Statement on p-Values: Context, Process, and Purpose.Am Stat. 2016 Apr 2;70(2):129–33. doi:10.1080/00031305.2016.1154108

20. Wasserstein RL, Schirm AL, Lazar NA. Moving to a World Beyond “plll<lll0.05”. Am Stat. 2019 Mar 29;73(sup1):1–19. doi:10.1080/00031305.2019.1583913

21. Teede H, Tay CT, Laven JSE. International Evidence-based Guideline for the Assessment and Management of Polycystic Ovary Syndrome 2023. Monash University; 2023.

22. NICE. Dyspepsia -proven GORD [Internet]. NICE; 2023. Available from: https://cks.nice.org.uk/topics/dyspepsia-proven-gord/

23. NICE. Diabetes -type 1 [Internet]. NICE; 2025. Available from: https://cks.nice.org.uk/topics/diabetes-type-1/

24. NICE. Diabetes -type 2 [Internet]. NICE; 2026. Available from: https://cks.nice.org.uk/topics/diabetes-type-2/

25. Wolf A, Dedman D, Campbell J, Booth H, Lunn D, Chapman J, et al. Data resource profile: Clinical Practice Research Datalink (CPRD) Aurum. Int J Epidemiol. 2019 Dec 1;48(6):1740–1740g. doi:10.1093/ije/dyz034

26. Rusch JA, van der Westhuizen DJ, Gill RS, Louw VJ. Diagnosing iron deficiency: Controversies and novel metrics. Best Pract Res Clin Anaesthesiol. 2023 Dec 1;37(4):451–67. doi:10.1016/j.bpa.2023.11.001

27. Kidney Disease: Improving, Global Outcomes (KDIGO) Anemia Work Group, Kidney Disease Improving Global Outcomes. KDIGO Clinical Practice Guideline for Anemia in Chronic Kidney Disease. J Int Soc Nephrol. 2012 Aug;2(4).

28. Dignass AU, Gasche C, Bettenworth D, Birgegård G, Danese S, Gisbert JP, et al. European Consensus on the Diagnosis and Management of Iron Deficiency and Anaemia in Inflammatory Bowel Diseases. J Crohns Colitis. 2015 Mar 1;9(3):211–22. doi:10.1093/ecco-jcc/jju009

