## Supplemental Tables and Figures for "Ferritin across long-term conditions in England: cross-sectional primary care study"

**Supplementary Tables & Figures**

**Supplementary Figure S1. Cohort flow diagram.**

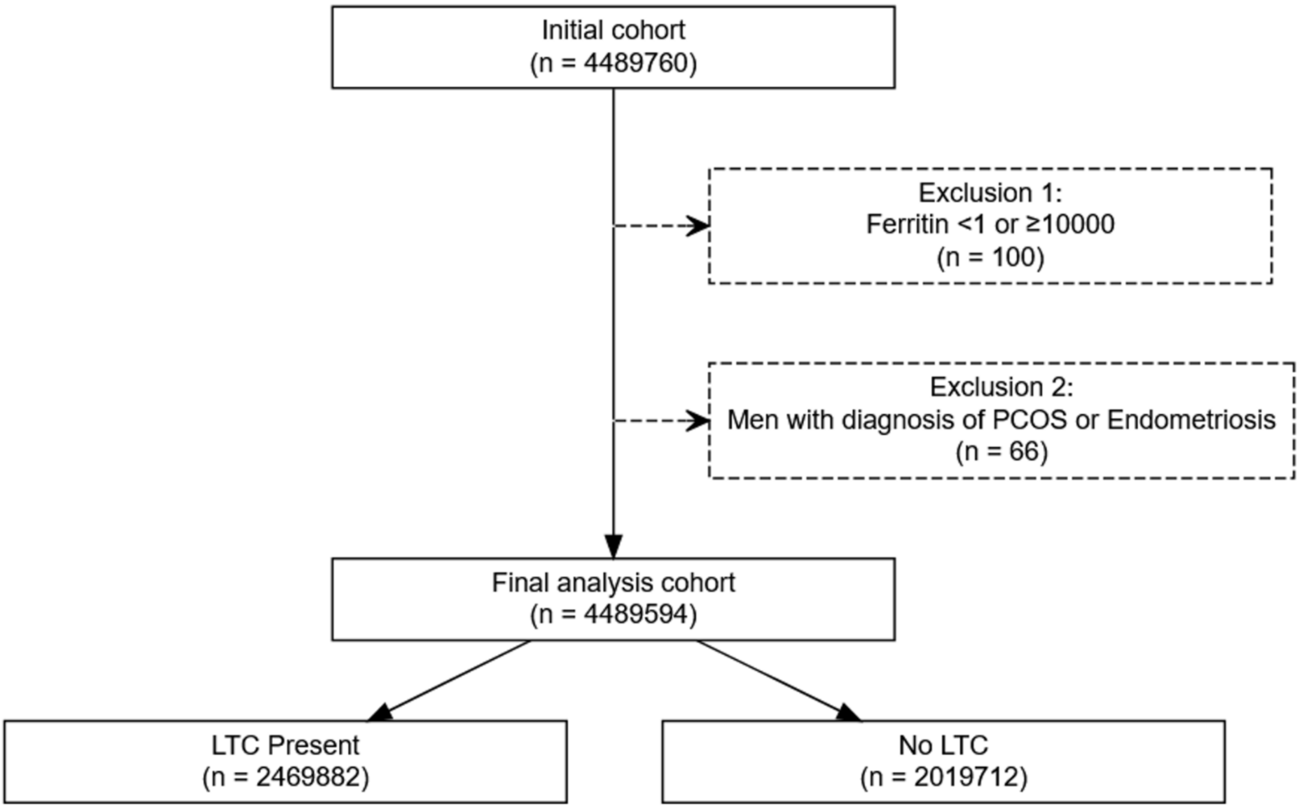

**Supplementary Table S1. LTC prevalence by sex.** Metrics are presented as N (%) stratified by sex.

| Cohort | Total | Male | Female |
|---|---|---|---|
| All Unique Individuals | 4489594 (100%) | 1565321 (34.9%) | 2924273 (65.1%) |
| No LTC | 2019712 (45%) | 673187 (33.3%) | 1346525 (66.7%) |
| LTC Present | 2469882 (55%) | 892134 (36.1%) | 1577748 (63.9%) |
| Bronchiectasis | 27547 (0.6%) | 10609 (38.5%) | 16938 (61.5%) |
| Cirrhosis | 12854 (0.3%) | 7087 (55.1%) | 5767 (44.9%) |
| CKD | 276533 (6.2%) | 108292 (39.2%) | 168241 (60.8%) |
| Coeliac | 26058 (0.6%) | 6838 (26.2%) | 19220 (73.8%) |
| COPD | 175355 (3.9%) | 83458 (47.6%) | 91897 (52.4%) |
| Crohn's Disease | 32857 (0.7%) | 12590 (38.3%) | 20267 (61.7%) |
| Dementia | 68478 (1.5%) | 25848 (37.7%) | 42630 (62.3%) |
| Diabetes (Type 1) | 30143 (0.7%) | 14379 (47.7%) | 15764 (52.3%) |
| Diabetes (Type 2) | 376343 (8.4%) | 192252 (51.1%) | 184091 (48.9%) |
| Endometriosis | 58565 (1.3%) | 0 (0%) | 58565 (100%) |
| GORD | 768966 (17.1%) | 285949 (37.2%) | 483017 (62.8%) |
| Heart Failure | 100481 (2.2%) | 54631 (54.4%) | 45850 (45.6%) |
| Multiple Sclerosis | 12927 (0.3%) | 3005 (23.2%) | 9922 (76.8%) |
| Obesity | 1229596 (27.4%) | 420145 (34.2%) | 809451 (65.8%) |
| Osteoarthritis | 711063 (15.8%) | 257918 (36.3%) | 453145 (63.7%) |
| Parkinson's Disease | 14847 (0.3%) | 8425 (56.7%) | 6422 (43.3%) |
| PCOS | 117455 (2.6%) | 0 (0%) | 117455 (100%) |
| Rheumatoid Arthritis | 138526 (3.1%) | 35839 (25.9%) | 102687 (74.1%) |
| Stroke | 127650 (2.8%) | 61083 (47.9%) | 66567 (52.1%) |
| Ulcerative Colitis | 39094 (0.9%) | 16936 (43.3%) | 22158 (56.7%) |

**Supplementary Table S2. Ferritin testing patterns by LTC and sex:** Metrics are presented as N (%) for counts, stratified by LTC and sex.

| Cohort | Number of Tests, n (%) | | |
|---|---|---|---|
| | Total | Male | Female |
| All Unique Individuals | 8774103 (100%) | 2815162 (32.1%) | 5958941 (67.9%) |
| Without LTMC | 3379560 (38.5%) | 992153 (29.4%) | 2387407 (70.6%) |
| With LTMC | 5394543 (61.5%) | 1823009 (33.8%) | 3571534 (66.2%) |
| Bronchiectasis | 67755 (0.8%) | 25014 (36.9%) | 42741 (63.1%) |
| Cirrhosis | 31463 (0.4%) | 16524 (52.5%) | 14939 (47.5%) |
| CKD | 697235 (7.9%) | 271464 (38.9%) | 425771 (61.1%) |
| Coeliac | 69290 (0.8%) | 16988 (24.5%) | 52302 (75.5%) |
| COPD | 414356 (4.7%) | 190800 (46%) | 223556 (54%) |
| Crohn's Disease | 81997 (0.9%) | 29729 (36.3%) | 52268 (63.7%) |
| Dementia | 126455 (1.4%) | 48440 (38.3%) | 78015 (61.7%) |
| Diabetes (Type 1) | 68987 (0.8%) | 30519 (44.2%) | 38468 (55.8%) |
| Diabetes (Type 2) | 964819 (11%) | 468706 (48.6%) | 496113 (51.4%) |
| Endometriosis | 129793 (1.5%) | 0 (0%) | 129793 (100%) |
| GORD | 1798181 (20.5%) | 605492 (33.7%) | 1192689 (66.3%) |
| Heart Failure | 245590 (2.8%) | 132156 (53.8%) | 113434 (46.2%) |
| Multiple Sclerosis | 27089 (0.3%) | 5635 (20.8%) | 21454 (79.2%) |
| Obesity | 2689242 (30.6%) | 836756 (31.1%) | 1852486 (68.9%) |
| Osteoarthritis | 1681104 (19.2%) | 585069 (34.8%) | 1096035 (65.2%) |
| Parkinson's Disease | 30464 (0.3%) | 16616 (54.5%) | 13848 (45.5%) |
| PCOS | 256273 (2.9%) | 0 (0%) | 256273 (100%) |
| Rheumatoid Arthritis | 345617 (3.9%) | 83417 (24.1%) | 262200 (75.9%) |
| Stroke | 290021 (3.3%) | 135427 (46.7%) | 154594 (53.3%) |
| Ulcerative Colitis | 93755 (1.1%) | 37976 (40.5%) | 55779 (59.5%) |

**Supplementary Table S3. Ferritin summary statistics by LTC and sex.** Median with 2.5th, 25th, 75th and 97.5th percentiles of serum ferritin (µg/L), stratified by LTC status and sex (men; women <50 years; women ≥50 years). Percentiles were calculated on the $\log_2$ scale and back-transformed to µg/L.

| Group | Condition | N | 2.5th percentile (µg/L) | 25th percentile (µg/L) | Median (µg/L) | 75th percentile (µg/L) | 97.5th percentile (µg/L) |
|---|---|---|---|---|---|---|---|
| Men (all ages) | No LTC | 673187 | 14 | 71 | 122 | 202 | 558 |
| Men (all ages) | LTC Present | 892134 | 9 | 50 | 109 | 204 | 606.9 |
| Men (all ages) | CKD | 108292 | 10 | 40 | 93 | 191 | 641.7 |
| Men (all ages) | Cirrhosis | 7087 | 8 | 30 | 84 | 202 | 916.8 |
| Men (all ages) | COPD | 83458 | 9 | 35.9 | 85 | 174 | 593 |
| Men (all ages) | Heart Failure | 54631 | 10 | 38 | 89 | 186.2 | 636 |
| Men (all ages) | Dementia | 25848 | 10 | 45 | 104 | 204 | 654 |
| Men (all ages) | Multiple Sclerosis | 3005 | 10 | 52 | 105.3 | 189 | 519.5 |
| Men (all ages) | Obesity | 420145 | 10 | 57 | 122 | 222 | 616 |
| Men (all ages) | Osteoarthritis | 257918 | 9.3 | 42.3 | 99 | 192.9 | 603 |
| Men (all ages) | Parkinson's Disease | 8425 | 10 | 43 | 97.3 | 187 | 570 |
| Men (all ages) | Bronchiectasis | 10609 | 9 | 36 | 87 | 176 | 580 |
| Men (all ages) | Diabetes (Type 1) | 14379 | 9 | 41 | 85 | 156 | 498.5 |
| Men (all ages) | Stroke | 61083 | 9 | 37 | 88 | 178 | 609 |
| Men (all ages) | Crohn's Disease | 12590 | 6 | 30 | 71 | 139 | 454.3 |
| Men (all ages) | Rheumatoid Arthritis | 35839 | 10 | 45 | 101 | 193 | 596 |
| Men (all ages) | GORD | 285949 | 9 | 44 | 99 | 186 | 553 |
| Men (all ages) | Coeliac | 6838 | 8 | 39 | 85 | 158.8 | 486 |
| Men (all ages) | Ulcerative Colitis | 16936 | 7 | 32 | 74 | 145 | 468.6 |
| Men (all ages) | Diabetes (Type 2) | 192252 | 8 | 29 | 70 | 152 | 557 |
| Women <50 | No LTC | 968335 | 4 | 15 | 29 | 53 | 153 |
| Women <50 | LTC Present | 583668 | 4.7 | 15 | 32 | 60 | 188.4 |
| Women <50 | CKD | 3067 | 6 | 20 | 45 | 104 | 601.9 |
| Women <50 | Cirrhosis | 947 | 5 | 15 | 37 | 82 | 678.7 |
| Women <50 | Endometriosis | 31366 | 5 | 17 | 35 | 65 | 192 |
| Women <50 | COPD | 4921 | 5 | 16 | 34 | 65 | 224 |
| Women <50 | Heart Failure | 1814 | 5 | 16 | 34 | 70 | 444.8 |
| Women <50 | Dementia | 1240 | 5 | 16 | 33 | 59 | 215.2 |
| Women <50 | Multiple Sclerosis | 2997 | 5 | 16 | 33 | 64 | 224.3 |
| Women <50 | Obesity | 371102 | 5 | 16 | 33 | 64 | 197 |
| Women <50 | PCOS | 89457 | 5 | 16 | 32 | 59 | 174 |
| Women <50 | Osteoarthritis | 21409 | 4 | 14 | 31.9 | 64 | 198 |
| Women <50 | Parkinson's Disease | 49 | 4.3 | 11 | 31 | 53 | 95.6 |
| Women <50 | Bronchiectasis | 1522 | 5 | 14.9 | 30 | 57 | 199.9 |
| Women <50 | Diabetes (Type 1) | 7502 | 4 | 14 | 30 | 57.9 | 204.4 |
| Women <50 | Stroke | 5183 | 4 | 14 | 30 | 59 | 218 |
| Women <50 | Crohn's Disease | 9700 | 4 | 14 | 29 | 55 | 188.5 |
| Women <50 | Rheumatoid Arthritis | 22182 | 4 | 14 | 29 | 57 | 186 |
| Women <50 | GORD | 150276 | 4 | 13 | 28 | 55 | 176 |
| Women <50 | Coeliac | 8736 | 4 | 13 | 27 | 52 | 158.4 |
| Women <50 | Ulcerative Colitis | 8360 | 4 | 13 | 27 | 52 | 176 |
| Women <50 | Diabetes (Type 2) | 18016 | 4 | 10 | 24 | 55.3 | 231.8 |
| Women ≥50 | No LTC | 378190 | 6 | 35.5 | 69.6 | 124 | 388 |
| Women ≥50 | LTC Present | 994080 | 7 | 27 | 61 | 119 | 395 |
| Women ≥50 | CKD | 165174 | 7.8 | 26 | 61 | 128 | 504 |
| Women ≥50 | Cirrhosis | 4820 | 7 | 22 | 55 | 137.1 | 783 |
| Women ≥50 | Endometriosis | 27199 | 7 | 33 | 65 | 116.3 | 333 |
| Women ≥50 | COPD | 86976 | 7 | 23 | 54 | 109 | 400 |
| Women ≥50 | Heart Failure | 44036 | 8 | 25 | 58 | 127 | 529 |
| Women ≥50 | Dementia | 41390 | 8 | 30 | 69 | 147 | 555.1 |
| Women ≥50 | Multiple Sclerosis | 6925 | 6 | 27 | 58 | 112 | 380.2 |
| Women ≥50 | Obesity | 438349 | 6 | 25 | 60 | 118 | 366 |
| Women ≥50 | PCOS | 27998 | 6 | 26 | 57 | 107 | 325 |
| Women ≥50 | Osteoarthritis | 431736 | 7 | 26.6 | 60 | 118 | 397 |
| Women ≥50 | Parkinson's Disease | 6373 | 7 | 26 | 59 | 119 | 436.3 |
| Women ≥50 | Bronchiectasis | 15416 | 7 | 24 | 55 | 109.4 | 380 |
| Women ≥50 | Diabetes (Type 1) | 8262 | 6 | 20 | 47 | 97 | 423.9 |
| Women ≥50 | Stroke | 61384 | 7 | 23 | 55 | 115 | 451 |
| Women ≥50 | Crohn's Disease | 10567 | 6 | 22 | 50 | 98.9 | 375 |
| Women ≥50 | Rheumatoid Arthritis | 80505 | 7 | 25.6 | 58 | 114 | 391 |
| Women ≥50 | GORD | 332741 | 6 | 24 | 54.9 | 108 | 356.1 |
| Women ≥50 | Coeliac | 10484 | 6 | 23 | 50 | 96 | 344.9 |
| Women ≥50 | Ulcerative Colitis | 13798 | 6 | 23 | 50.3 | 97 | 370 |
| Women ≥50 | Diabetes (Type 2) | 166075 | 6 | 17 | 41 | 93 | 397 |

**Supplementary Table S4. Threshold-defined low ferritin counts/percentages by sex and LTC status at <15, <30 and <70 µg/L with binomial (Wilson) 95% CIs.**

| Group | LTC Status | N | Low Ferritin Prevalence, <15 µg/L (%, 95% CI) | Low Ferritin Prevalence, <30 µg/L (%, 95% CI) | Low Ferritin Prevalence, <70 µg/L (%, 95% CI) |
|---|---|---|---|---|---|
| Men (all ages) | LTC Present | 461635 | 4.3 (4.3-4.4) | 10.7 (10.6-10.8) | 27.7 (27.6-27.8) |
| Men (all ages) | No LTC | 673187 | 2.6 (2.5-2.6) | 6.7 (6.7-6.8) | — |
| Women <50 | LTC Present | 440822 | 23.3 (23.2-23.5) | 47 (46.9-47.2) | 80.1 (79.9-80.2) |
| Women <50 | No LTC | 968335 | 24.5 (24.4-24.6) | 50.3 (50.2-50.4) | — |
| Women ≥50 | LTC Present | 423398 | 10.3 (10.2-10.3) | 22.5 (22.4-22.6) | 50.8 (50.6-50.9) |
| Women ≥50 | No LTC | 378190 | 9.3 (9.3-9.4) | 20.4 (20.3-20.5) | — |

**Supplementary Table S5. Prevalence of threshold-defined low ferritin by LTC and sex with binomial (Wilson) 95% CIs.**

| Group | Condition | N | Threshold-define Low Ferritin (%, 95% CI) |
|---|---|---|---|
| Men (all ages) | No LTC | 673187 | 2.6 (2.5-2.6) |
| Men (all ages) | LTC Present | 892134 | 34 (33.9-34.1) |
| Men (all ages) | CKD | 108292 | 40.4 (40.1-40.6) |
| Men (all ages) | Cirrhosis | 7087 | 45.1 (44-46.3) |
| Men (all ages) | Heart Failure | 54631 | 41.8 (41.4-42.2) |
| Men (all ages) | Endometriosis | 0 | 0 (0-0) |
| Men (all ages) | COPD | 83458 | 43.1 (42.8-43.5) |
| Men (all ages) | Obesity | 420145 | 30.1 (30-30.3) |
| Men (all ages) | Multiple Sclerosis | 3005 | 34 (32.3-35.7) |
| Men (all ages) | Osteoarthritis | 257918 | 38.2 (38-38.4) |
| Men (all ages) | PCOS | 0 | 0 (0-0) |
| Men (all ages) | Dementia | 25848 | 36.7 (36.2-37.3) |
| Men (all ages) | Crohn's Disease | 12590 | 49.5 (48.6-50.3) |
| Men (all ages) | Diabetes (Type 1) | 14379 | 42.1 (41.3-42.9) |
| Men (all ages) | Rheumatoid Arthritis | 35839 | 37 (36.5-37.5) |
| Men (all ages) | Stroke | 61083 | 42.4 (42-42.8) |
| Men (all ages) | Bronchiectasis | 10609 | 42.8 (41.9-43.7) |
| Men (all ages) | GORD | 285949 | 37.4 (37.2-37.6) |
| Men (all ages) | Ulcerative Colitis | 16936 | 47.9 (47.1-48.6) |
| Men (all ages) | Parkinson's Disease | 8425 | 38.4 (37.4-39.4) |
| Men (all ages) | Coeliac | 6838 | 42.6 (41.4-43.8) |
| Men (all ages) | Diabetes (Type 2) | 192252 | 49.8 (49.6-50) |
| Women <50 | No LTC | 968335 | 24.5 (24.4-24.6) |
| Women <50 | LTC Present | 583668 | 79.7 (79.6-79.8) |
| Women <50 | CKD | 3067 | 64.3 (62.6-66) |
| Women <50 | Cirrhosis | 947 | 69.7 (66.8-72.6) |
| Women <50 | Heart Failure | 1814 | 74.9 (72.9-76.9) |
| Women <50 | Endometriosis | 31366 | 77.7 (77.2-78.1) |
| Women <50 | COPD | 4921 | 77.4 (76.2-78.5) |
| Women <50 | Obesity | 371102 | 77.8 (77.6-77.9) |
| Women <50 | Multiple Sclerosis | 2997 | 77.7 (76.2-79.2) |
| Women <50 | Osteoarthritis | 21409 | 77.9 (77.3-78.4) |
| Women <50 | PCOS | 89457 | 80.9 (80.7-81.2) |
| Women <50 | Dementia | 1240 | 80.2 (77.9-82.4) |
| Women <50 | Crohn's Disease | 9700 | 82 (81.2-82.8) |
| Women <50 | Diabetes (Type 1) | 7502 | 80.8 (79.9-81.7) |
| Women <50 | Rheumatoid Arthritis | 22182 | 81.3 (80.8-81.8) |
| Women <50 | Stroke | 5183 | 79.9 (78.8-81) |
| Women <50 | Bronchiectasis | 1522 | 80.9 (78.9-82.9) |
| Women <50 | GORD | 150276 | 82.5 (82.3-82.7) |
| Women <50 | Ulcerative Colitis | 8360 | 83.5 (82.7-84.3) |
| Women <50 | Parkinson's Disease | 49 | 81.6 (70.8-92.5) |
| Women <50 | Coeliac | 8736 | 84.7 (83.9-85.5) |
| Women <50 | Diabetes (Type 2) | 18016 | 80.5 (79.9-81.1) |
| Women ≥50 | No LTC | 378190 | 9.3 (9.3-9.4) |
| Women ≥50 | LTC Present | 994080 | 54.8 (54.7-54.9) |
| Women ≥50 | CKD | 165174 | 54.4 (54.1-54.6) |
| Women ≥50 | Cirrhosis | 4820 | 56.5 (55.1-57.9) |
| Women ≥50 | Heart Failure | 44036 | 55.8 (55.4-56.3) |
| Women ≥50 | Endometriosis | 27199 | 52.8 (52.2-53.4) |
| Women ≥50 | COPD | 86976 | 59.3 (59-59.7) |
| Women ≥50 | Obesity | 438349 | 55.2 (55.1-55.4) |
| Women ≥50 | Multiple Sclerosis | 6925 | 56.5 (55.3-57.6) |
| Women ≥50 | Osteoarthritis | 431736 | 55.5 (55.4-55.7) |
| Women ≥50 | PCOS | 27998 | 57.8 (57.2-58.4) |
| Women ≥50 | Dementia | 41390 | 50.2 (49.7-50.6) |
| Women ≥50 | Crohn's Disease | 10567 | 62.7 (61.8-63.7) |
| Women ≥50 | Diabetes (Type 1) | 8262 | 63.5 (62.4-64.5) |
| Women ≥50 | Rheumatoid Arthritis | 80505 | 57 (56.7-57.3) |
| Women ≥50 | Stroke | 61384 | 58.2 (57.8-58.6) |
| Women ≥50 | Bronchiectasis | 15416 | 58.5 (57.7-59.3) |
| Women ≥50 | GORD | 332741 | 59 (58.9-59.2) |
| Women ≥50 | Ulcerative Colitis | 13798 | 62.4 (61.6-63.2) |
| Women ≥50 | Parkinson's Disease | 6373 | 56 (54.7-57.2) |
| Women ≥50 | Coeliac | 10484 | 62.7 (61.8-63.7) |
| Women ≥50 | Diabetes (Type 2) | 166075 | 66.6 (66.3-66.8) |

**Supplementary Table S6a. Multimorbidity (MLTC) vs single LTC: Geometric mean ferritin (µg/L) with 95% CI, stratified by sex and age group.** Geometric means were derived from $log_2$-transformed ferritin values and back-transformed to µg/L. Single LTC = exactly one LTC; Multiple LTCs = two or more LTCs. Men (all ages)

| Condition | Geometric Mean Ferritin (95% CI) Single LTC | N (Single LTC) | Geometric Mean Ferritin (95% CI) Multiple LTCs | N (Multiple LTCs) |
|---|---|---|---|---|
| Bronchiectasis | 99.1 (93.9–104.5) | 1,394 | 76.6 (74.9–78.4) | 9,215 |
| Cirrhosis | 105.7 (98.9–113) | 1,444 | 75 (72.5–77.5) | 5,643 |
| CKD | 105.3 (103.6–107.1) | 15,947 | 84 (83.4–84.6) | 92,345 |
| Coeliac | 83.6 (80.6–86.8) | 2,717 | 71.8 (69.5–74.2) | 4,121 |
| COPD | 99.7 (98.1–101.4) | 15,544 | 74.2 (73.6–74.9) | 67,914 |
| Crohn's Disease | 61 (58.9–63.1) | 4,076 | 63.5 (62–65.1) | 8,514 |
| Dementia | 113 (109.7–116.4) | 4,562 | 90.9 (89.6–92.2) | 21,286 |
| Diabetes (Type 1) | 89.5 (87–92.1) | 3,877 | 73.9 (72.4–75.4) | 10,502 |
| Diabetes (Type 2) | 69.7 (68.9–70.6) | 33,605 | 66.3 (66–66.7) | 158,647 |
| Endometriosis | NA | 0 | NA | 0 |
| GORD | 102.2 (101.6–102.8) | 102,256 | 79.8 (79.4–80.2) | 183,693 |
| Heart Failure | 101.4 (98.8–104.2) | 6,120 | 82.1 (81.3–83) | 48,511 |
| Multiple Sclerosis | 107 (102–112.2) | 1,352 | 84.7 (80.4–89.3) | 1,653 |
| Obesity | 135.4 (134.8–136) | 183,279 | 86.7 (86.3–87.1) | 236,866 |
| Osteoarthritis | 108.2 (107.3–109.1) | 60,493 | 83.1 (82.7–83.5) | 197,425 |
| Parkinson's Disease | 101.8 (97.2–106.5) | 1,786 | 84.3 (82.1–86.5) | 6,639 |
| PCOS | NA | 0 | NA | 0 |
| Rheumatoid Arthritis | 106 (103.8–108.4) | 7,641 | 86.2 (85.1–87.3) | 28,198 |
| Stroke | 95.3 (93.3–97.3) | 10,661 | 77.5 (76.7–78.2) | 50,422 |
| Ulcerative Colitis | 66.5 (64.5–68.6) | 4,881 | 66.1 (64.8–67.4) | 12,055 |

**Supplementary Table S6a (continued).** Multimorbidity (MLTC) vs single LTC: Geometric mean ferritin (µg/L) with 95% CI. Women aged <50 years.

| Condition | Geometric Mean Ferritin (95% CI) Single LTC | N (Single LTC) | Geometric Mean Ferritin (95% CI) Multiple LTCs | N (Multiple LTCs) |
|---|---|---|---|---|
| Bronchiectasis | 27.4 (25.6–29.4) | 716 | 31.1 (29–33.4) | 806 |
| Cirrhosis | 41.3 (36.1–47.1) | 403 | 37.3 (33.7–41.2) | 544 |
| CKD | 46.6 (43.3–50.2) | 1,044 | 49.1 (46.5–51.8) | 2,023 |
| Coeliac | 26.1 (25.4–26.7) | 5,381 | 25.9 (25.1–26.8) | 3,355 |
| COPD | 33.4 (31.9–34.9) | 1,848 | 32.4 (31.2–33.6) | 3,073 |
| Crohn's Disease | 27.7 (26.8–28.6) | 3,815 | 28.1 (27.4–28.8) | 5,885 |
| Dementia | 30.4 (28.4–32.7) | 673 | 32.1 (29.6–34.9) | 567 |
| Diabetes (Type 1) | 29.3 (28.3–30.3) | 3,128 | 28.9 (28–29.8) | 4,374 |
| Diabetes (Type 2) | 21.9 (21–22.8) | 2,950 | 25.7 (25.2–26.1) | 15,066 |
| Endometriosis | 32 (31.5–32.5) | 15,304 | 33.7 (33.2–34.2) | 16,062 |
| GORD | 26.3 (26.1–26.5) | 80,097 | 28.4 (28.1–28.6) | 70,179 |
| Heart Failure | 32.2 (29.6–35) | 639 | 36.5 (34.1–39.1) | 1,175 |
| Multiple Sclerosis | 31.9 (30.5–33.4) | 1,623 | 32.8 (31–34.7) | 1,374 |
| Obesity | 32.1 (32–32.2) | 258,899 | 30.6 (30.4–30.7) | 112,203 |
| Osteoarthritis | 29.6 (28.9–30.3) | 7,119 | 30.3 (29.8–30.9) | 14,290 |
| Parkinson's Disease | 21.8 (15.4–30.9) | 24 | 33.5 (21.3–52.9) | 25 |
| PCOS | 29.4 (29.2–29.7) | 41,714 | 31.2 (30.9–31.4) | 47,743 |
| Rheumatoid Arthritis | 27.4 (26.9–27.9) | 9,672 | 29.1 (28.6–29.6) | 12,510 |
| Stroke | 27.9 (26.7–29) | 2,451 | 29.9 (28.8–31.2) | 2,732 |
| Ulcerative Colitis | 25.4 (24.6–26.3) | 3,322 | 27.2 (26.5–28) | 5,038 |

**Supplementary Table S6a (continued).** Multimorbidity (MLTC) vs single LTC: Geometric mean ferritin (µg/L) with 95% CI. Women aged ≥50 years.

| Condition | Geometric Mean Ferritin (95% CI) Single LTC | N (Single LTC) | Geometric Mean Ferritin (95% CI) Multiple LTCs | N (Multiple LTCs) |
|---|---|---|---|---|
| Bronchiectasis | 64.8 (62.2–67.5) | 2,057 | 50.4 (49.5–51.3) | 13,359 |
| Cirrhosis | 77.3 (70.4–84.8) | 777 | 54.2 (52.2–56.3) | 4,043 |
| CKD | 70.8 (69.8–71.8) | 21,449 | 57.4 (57.1–57.8) | 143,725 |
| Coeliac | 47.3 (45.6–49) | 3,140 | 46.7 (45.6–47.9) | 7,344 |
| COPD | 63.1 (62–64.2) | 13,523 | 49 (48.6–49.4) | 73,453 |
| Crohn's Disease | 51.2 (48.8–53.9) | 1,866 | 46.1 (45.1–47.2) | 8,701 |
| Dementia | 74 (72.1–75.9) | 7,004 | 65.5 (64.7–66.3) | 34,386 |
| Diabetes (Type 1) | 47.8 (44.7–51.1) | 993 | 45.8 (44.6–47) | 7,269 |
| Diabetes (Type 2) | 42.6 (41.9–43.3) | 17,778 | 41.5 (41.2–41.7) | 148,297 |
| Endometriosis | 62.6 (61.3–63.9) | 8,017 | 57.4 (56.6–58.2) | 19,182 |
| GORD | 57.1 (56.7–57.5) | 76,491 | 48.6 (48.4–48.8) | 256,250 |
| Heart Failure | 67.4 (65–70) | 3,300 | 57.6 (57–58.3) | 40,736 |
| Multiple Sclerosis | 58.6 (56.4–61) | 2,542 | 51.2 (49.6–52.9) | 4,383 |
| Obesity | 63.7 (63.4–64.1) | 134,883 | 50.2 (50–50.4) | 303,466 |
| Osteoarthritis | 66.1 (65.7–66.5) | 96,534 | 53.2 (53–53.4) | 335,202 |
| Parkinson's Disease | 65.3 (61.7–69.1) | 1,206 | 55 (53.4–56.7) | 5,167 |
| PCOS | 54.2 (52.9–55.5) | 6,995 | 50.3 (49.5–51) | 21,003 |
| Rheumatoid Arthritis | 60.1 (59.1–61.1) | 13,794 | 52.8 (52.4–53.3) | 66,711 |
| Stroke | 59.2 (57.8–60.5) | 8,376 | 52 (51.5–52.5) | 53,008 |
| Ulcerative Colitis | 52.4 (50.4–54.5) | 2,673 | 46.2 (45.3–47.1) | 11,125 |

**Supplementary Table S6b.** Multimorbidity (MLTC) vs single LTC: Threshold-defined low ferritin prevalence (<70 µg/L, %) with binomial (Wilson) 95% CI, stratified by sex and age group. Single LTC = exactly one LTC; Multiple LTCs = two or more LTCs. Men (all ages).

| Condition | Threshold-defined low ferritin % (95% CI) Single LTC | N (Single LTC) | Threshold-defined low ferritin % (95% CI) Multiple LTCs | N (Multiple LTCs) |
|---|---|---|---|---|
| Bronchiectasis | 31.4 (29–33.9) | 1,394 | 44.5 (43.5–45.5) | 9,215 |
| Cirrhosis | 35.6 (33.1–38.1) | 1,444 | 47.5 (46.2–48.8) | 5,643 |
| CKD | 31.7 (31–32.5) | 15,947 | 41.8 (41.5–42.2) | 92,345 |
| Coeliac | 37.1 (35.2–38.9) | 2,717 | 46.3 (44.8–47.8) | 4,121 |
| COPD | 33 (32.2–33.7) | 15,544 | 45.5 (45.1–45.8) | 67,914 |
| Crohn's Disease | 49.5 (48–51.1) | 4,076 | 49.4 (48.4–50.5) | 8,514 |
| Dementia | 28.8 (27.5–30.1) | 4,562 | 38.4 (37.8–39.1) | 21,286 |
| Diabetes (Type 1) | 34 (32.5–35.5) | 3,877 | 45.1 (44.1–46) | 10,502 |
| Diabetes (Type 2) | 48.5 (48–49.1) | 33,605 | 50.1 (49.8–50.3) | 158,647 |
| Endometriosis | NA | 0 | NA | 0 |
| GORD | 29.6 (29.3–29.9) | 102,256 | 41.8 (41.5–42) | 183,693 |
| Heart Failure | 33.2 (32–34.3) | 6,120 | 42.9 (42.4–43.3) | 48,511 |
| Multiple Sclerosis | 28.3 (25.9–30.7) | 1,352 | 38.8 (36.4–41.1) | 1,653 |
| Obesity | 19.5 (19.3–19.7) | 183,279 | 38.3 (38.1–38.5) | 236,866 |
| Osteoarthritis | 29.3 (28.9–29.7) | 60,493 | 41 (40.7–41.2) | 197,425 |
| Parkinson's Disease | 30.5 (28.3–32.6) | 1,786 | 40.5 (39.4–41.7) | 6,639 |
| PCOS | NA | 0 | NA | 0 |
| Rheumatoid Arthritis | 28.6 (27.6–29.6) | 7,641 | 39.3 (38.7–39.8) | 28,198 |
| Stroke | 33.8 (32.9–34.7) | 10,661 | 44.2 (43.7–44.6) | 50,422 |
| Ulcerative Colitis | 47 (45.6–48.4) | 4,881 | 48.2 (47.3–49.1) | 12,055 |

**Supplementary Table S6b (continued).** Multimorbidity (MLTC) vs single LTC: Threshold-defined low ferritin prevalence (<70 µg/L, %) with binomial (Wilson) 95% CI. Women aged <50 years.

| Condition | Threshold-defined low ferritin % (95% CI) Single LTC | N (Single LTC) | Threshold-defined low ferritin % (95% CI) Multiple LTCs | N (Multiple LTCs) |
|---|---|---|---|---|
| Bronchiectasis | 82.8 (80.1–85.6) | 716 | 79.2 (76.4–82) | 806 |
| Cirrhosis | 66 (61.4–70.6) | 403 | 72.4 (68.7–76.2) | 544 |
| CKD | 67.3 (64.5–70.2) | 1,044 | 62.7 (60.6–64.8) | 2,023 |
| Coeliac | 85.7 (84.8–86.6) | 5,381 | 83.1 (81.8–84.4) | 3,355 |
| COPD | 78.1 (76.2–80) | 1,848 | 76.9 (75.4–78.4) | 3,073 |
| Crohn's Disease | 82.8 (81.6–84) | 3,815 | 81.5 (80.5–82.5) | 5,885 |
| Dementia | 82.2 (79.3–85.1) | 673 | 77.8 (74.4–81.2) | 567 |
| Diabetes (Type 1) | 81.9 (80.6–83.3) | 3,128 | 80 (78.8–81.2) | 4,374 |
| Diabetes (Type 2) | 84.7 (83.4–86) | 2,950 | 79.7 (79.1–80.3) | 15,066 |
| Endometriosis | 79.4 (78.8–80.1) | 15,304 | 76 (75.3–76.7) | 16,062 |
| GORD | 84.5 (84.3–84.8) | 80,097 | 80.2 (79.9–80.5) | 70,179 |
| Heart Failure | 79.3 (76.2–82.5) | 639 | 72.5 (70–75.1) | 1,175 |
| Multiple Sclerosis | 79.1 (77.1–81) | 1,623 | 76.1 (73.9–78.4) | 1,374 |
| Obesity | 77.8 (77.7–78) | 258,899 | 77.7 (77.5–77.9) | 112,203 |
| Osteoarthritis | 79.9 (78.9–80.8) | 7,119 | 76.9 (76.2–77.6) | 14,290 |
| Parkinson's Disease | 87.5 (74.3–100.7) | 24 | 76 (59.3–92.7) | 25 |
| PCOS | 83.4 (83.1–83.8) | 41,714 | 78.8 (78.4–79.1) | 47,743 |
| Rheumatoid Arthritis | 83.5 (82.7–84.2) | 9,672 | 79.6 (78.9–80.3) | 12,510 |
| Stroke | 82.5 (81–84) | 2,451 | 77.6 (76–79.1) | 2,732 |
| Ulcerative Colitis | 85 (83.8–86.3) | 3,322 | 82.5 (81.5–83.6) | 5,038 |

**Supplementary Table S6b (continued).** Multimorbidity (MLTC) vs single LTC: Threshold-defined low ferritin deficiency prevalence (<70 µg/L, %) with binomial (Wilson) 95% CI. Women aged ≥50 years.

| Condition | Threshold-defined low ferritin % (95% CI) Single LTC | N (Single LTC) | Threshold-defined low ferritin % (95% CI) Multiple LTCs | N (Multiple LTCs) |
|---|---|---|---|---|
| Bronchiectasis | 50.4 (48.2–52.5) | 2,057 | 59.8 (58.9–60.6) | 13,359 |
| Cirrhosis | 47 (43.5–50.5) | 777 | 58.3 (56.8–59.9) | 4,043 |
| CKD | 46.6 (46–47.3) | 21,449 | 55.5 (55.3–55.8) | 143,725 |
| Coeliac | 63 (61.3–64.7) | 3,140 | 62.6 (61.5–63.7) | 7,344 |
| COPD | 51.3 (50.4–52.1) | 13,523 | 60.8 (60.5–61.2) | 73,453 |
| Crohn's Disease | 59.6 (57.4–61.9) | 1,866 | 63.4 (62.4–64.4) | 8,701 |
| Dementia | 45.9 (44.7–47) | 7,004 | 51 (50.5–51.6) | 34,386 |
| Diabetes (Type 1) | 60.4 (57.4–63.5) | 993 | 63.9 (62.8–65) | 7,269 |
| Diabetes (Type 2) | 65.8 (65.1–66.5) | 17,778 | 66.7 (66.4–66.9) | 148,297 |
| Endometriosis | 50.9 (49.8–52) | 8,017 | 53.6 (52.9–54.3) | 19,182 |
| GORD | 54 (53.7–54.4) | 76,491 | 60.5 (60.3–60.7) | 256,250 |
| Heart Failure | 50.5 (48.7–52.2) | 3,300 | 56.3 (55.8–56.7) | 40,736 |
| Multiple Sclerosis | 55 (53.1–57) | 2,542 | 57.3 (55.8–58.8) | 4,383 |
| Obesity | 48 (47.8–48.3) | 134,883 | 58.4 (58.3–58.6) | 303,466 |
| Osteoarthritis | 48.7 (48.4–49) | 96,534 | 57.5 (57.3–57.7) | 335,202 |
| Parkinson's Disease | 49.6 (46.8–52.4) | 1,206 | 57.4 (56.1–58.8) | 5,167 |
| PCOS | 55.9 (54.7–57) | 6,995 | 58.4 (57.8–59.1) | 21,003 |
| Rheumatoid Arthritis | 52.5 (51.6–53.3) | 13,794 | 57.9 (57.6–58.3) | 66,711 |
| Stroke | 53.6 (52.6–54.7) | 8,376 | 59 (58.5–59.4) | 53,008 |
| Ulcerative Colitis | 58.1 (56.2–59.9) | 2,673 | 63.5 (62.6–64.4) | 11,125 |
